# Wearables and smartphones for modifiable risk factors in metabolic health: a scoping review protocol

**DOI:** 10.1101/2024.04.15.24305819

**Authors:** Victoria Brügger, Tobias Kowatsch, Mia Jovanova

**Affiliations:** School of Medicine, University of St. Gallen, St. Gallen, Switzerland; Institute for Implementation Science in Health Care, University of Zurich, Zurich, Switzerland; Centre for Digital Health Interventions, Department of Management, Technology and Economics, ETH Zürich, Zurich, Switzerland

**Keywords:** wearables, smartphones, mHealth, metabolic diseases, risk factors, lifestyle, physiological, metabolic health

## Abstract

**Background:** Metabolic diseases, such as cardiovascular diseases and diabetes, contribute significantly to global mortality and disability. Wearable devices and smartphones increasingly track physiological and lifestyle risk factors and can improve the management of metabolic diseases. However, the absence of clear guidelines for deriving meaningful signals from these devices often hampers cross-study comparisons.

**Objective:** Thus, this scoping review protocol aims to systematically overview the current empirical literature on how wearables and smartphones are used to measure modifiable risk factors associated with metabolic diseases.

**Methods:** We will conduct a scoping review to overview how wearables and smartphones measure modifiable risk factors related to metabolic diseases. We will search six databases (Scopus, Web of Science, ScienceDirect, PubMed, ACM Digital Library, and IEEE Xplore) from 2019 to 2024, with search terms related to wearables, smartphones, and modifiable risk factors associated with metabolic diseases. We will apply the PRISMA-ScR (Preferred Reporting Items for Systematic Reviews and Meta-Analyses extension for Scoping Reviews) and Arksey and O’Malley’s scoping review methodology. Eligible studies will use smartphones and/or wearables (worn on the wrist, finger, arm, hip, and chest) to track physiological and/or lifestyle factors related to metabolic diseases. Two reviewers will independently screen articles for inclusion. Data will be extracted using a standardized form, and the findings will be synthesized and reported qualitatively and quantitatively.

**Results:** The study is expected to identify potential gaps in measuring modifiable risk factors in current digital metabolic health research. Results are expected to inform more standardized guidelines on wearable and smartphone-based measurements to aid cross-study comparison. The final report is planned for submission to an indexed journal.

**Conclusions:** This review is among the first to systematically overview the current landscape on how wearables and smartphones are used to measure modifiable risk factors associated with metabolic diseases.

## Introduction

Non-communicable diseases (NCD) globally lead to 41 million deaths per year and are estimated to cost, on average, more than US$ 2 trillion per year [1,2]. A large portion of NCD-related burden is attributed to a growing prevalence of metabolic diseases, namely type 2 diabetes, hypertension, hyperlipidemia, obesity, and, more recently, non-alcoholic fatty liver disease [3,4]. Metabolic diseases are projected to increase at an alarming rate, such that the number of people with diabetes alone is expected to double from 529 million in 2021 to 1.3 billion in 2050, with projected health expenditure above US$ 1054 billion by 2045 [5].

A large body of research has shown that metabolic diseases, e.g., type 2 diabetes, are influenced by a network of modifiable factors. These include lifestyle determinants (i.e., nutrition, physical activity, sleep, stress, and substance abuse) and physiological markers (i.e., blood sugar, triglycerides, and HDL cholesterol) [6–10]. Following a complex systems perspective, modifiable factors interact dynamically [11] and together shape disease outcomes over time, with up to 70% of cardiovascular disease cases and mortality attributed to modifiable risk factors [12] [9].

In parallel, wearables (i.e., devices worn on the wrist, finger, arm, and chest) and smartphones are increasingly used to track modifiable risk factors in daily life with improved precision and accuracy [13]. These devices offer key advantages over lab-based visits, such as continuous and person-specific data collection in (near) real-time. For instance, smartwatches can track physical activity and sleep patterns with minimal burden [15]. Further, mobile apps can detect nutrition via image-based food recognition [14], ecologic momentary assessment, or short surveys [16] in free-living conditions.

These high-dimensional, longitudinal data can trigger personalized lifestyle interventions by deploying recommendations on nutrition, sleep, and physical activity [15,16]. By enabling remote monitoring, wearables, and smartphones can improve access to care and reduce the costs of metabolic disease management [17,18] compared to standard lab-based clinical approaches [22].

While studies increasingly demonstrate the promise of wearables for tracking and managing metabolic diseases [19], key gaps remain in the newly evolving digital metabolic health sphere. Here, we focus on two. First, recent perspectives highlight the importance of tracking multi-modal vs. single risk factors [12] for a more comprehensive lens into an individual’s metabolic health profile [20]. However, the extent to which different studies focus on one vs. more risk factors in digital metabolic health contexts remains unclear. Second, alongside the proliferation of studies using wearables and smartphones, there are concerns regarding the comparability of data even when measuring the same risk factors [21]. Specifically, there is growing evidence of incommensurability, with different researchers employing different operationalizations (or units) when studying the same risk factor (i.e., physical inactivity), thus making direct cross-study comparisons challenging. This heterogeneity makes it difficult to track between-study effects [22] and presents a barrier to building cumulative and generalizable knowledge in this young and rapidly evolving field [23]. Motivated by these gaps, scoping the landscape of what risk factors are measured, and how, is essential for advancing the use of wearables and smartphones in digital metabolic health research.

Thus far, recent work has begun to overview the use of different wearable technologies in cardiometabolic diseases [23] and the role of digital health technologies in metabolic disorders among older adults, more broadly [25]. However, to our knowledge, no studies have specifically focused on the landscape of modifiable risk factors. Thus, we aim to address the following questions:

1. Which modifiable risk factors are most often studied in wearable and smartphone-based metabolic health research?
2. To what extent are measures of modifiable risk factors consistent across studies, particularly in measurement methods?

Gaining a comprehensive understanding of the current risk factor landscape is crucial to inform more consistent measurement guidelines. Given the broad nature of the inquiry and the emerging status of the field, we deemed a scoping review the most appropriate method for investigating these research questions.

## Methods

This scoping review follows the five-stage approach by Arksey and O’Malley [24]: (1) identifying the research question, (2) identifying relevant literature, (3) study selection, (4) charting the data, (5) collating, summarizing and reporting the articles. To guarantee adherence to guidelines, we adopt the PRISMA-ScR (Preferred Reporting Items for Systematic Reviews and Meta-Analyses extension for Scoping Reviews) checklist [25].

### Stage 1: identifying the research question

Motivated by gaps in prior work [22,26] and a lack of overview in the field, this scoping review aims to systematically scope current research regarding (1) which modifiable risk factors are most often studied (vs. understudied) in wearable and smartphone-based metabolic health research, and (2) to what extent are measures of modifiable risk factors consistent across studies in terms of measurement methods and units.

### Stage 2: identifying relevant studies

Drawing on the most recent reviews in the wearable, digital health, and metabolic health domains [22,26], we identified key search terms capturing wearables and/or smartphones and prominent modifiable risk factors associated with metabolic diseases [6]. See Appendix Table S1 for a full list of key search terms. We will search the following six major databases: Scopus, Web of Science, ScienceDirect, PubMed, and IEEE Xplore. The articles identified by the search will be imported into the collaborative, systematic review tool Rayyan [27].

### Stage 3: study selection: inclusion and exclusion criteria

Eligible studies will be peer-reviewed, written in English, published in 2019 and after (to reflect the most current digital metabolic health literature [28]); and will include a full-text version. In cases where no full text is available, authors will be contacted with a waiting period of 7 days between each contact before study exclusion. Given our research aims, eligible studies will (1) measure lifestyle and physiological factors relevant to metabolic health using (2) smartphones and/or wearables, which are worn on the wrist, finger, arm, and chest, and (3) analyze the generated wearable/smartphone data for outcome assessment. Qualitative studies will be excluded. Building on prior work [28], we will focus only on wearable devices that cost <€500 (US $570) per device hardware to ensure that wearables are available commercially, intended for consumers, and potentially allow for scalability. We will include membership-based devices.

### Step 4: charting the data

For each study deemed eligible and included in the review, relevant information will be extracted and documented using a standardized data charting form. Two reviewers will manually chart the extracted information, and a third reviewer will resolve discrepancies. See Table 2 for the data charting form.

**Table 1.**
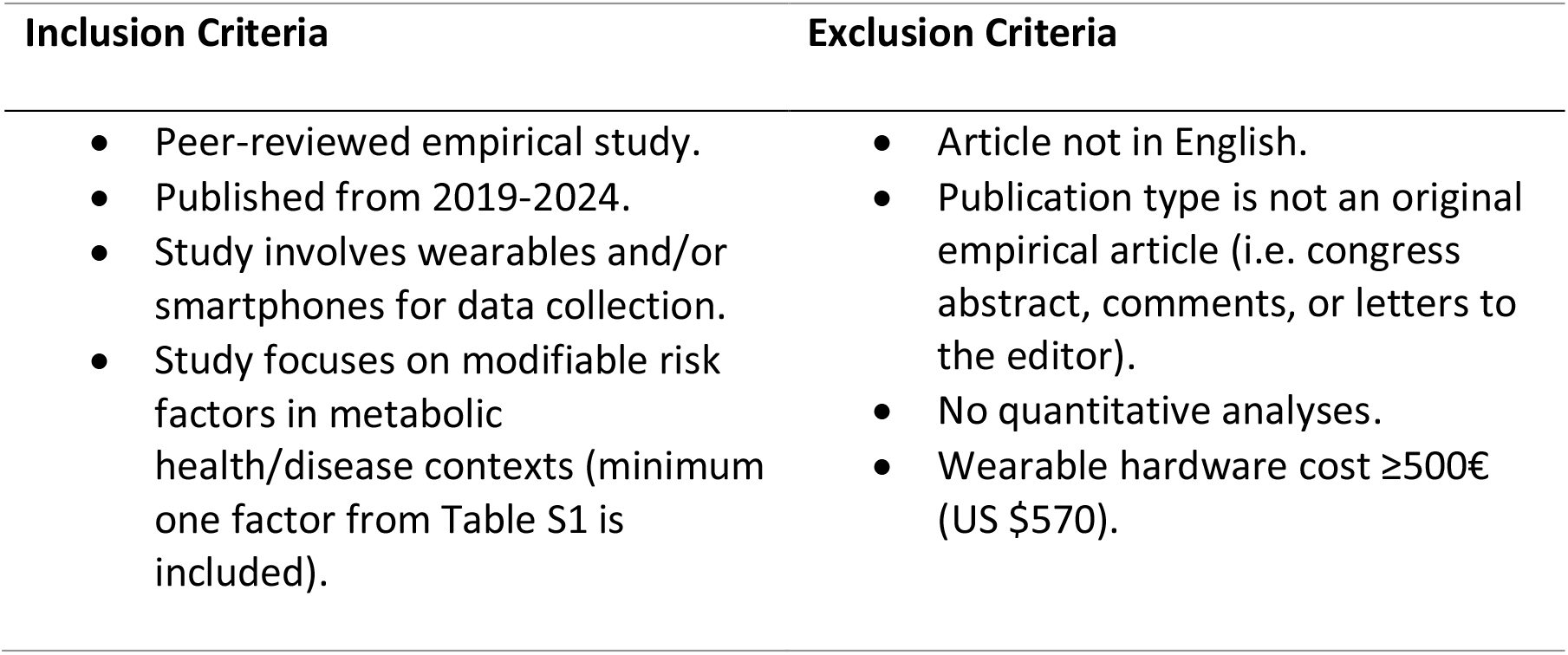
Study inclusion and exclusion criteria.

| Inclusion Criteria | Exclusion Criteria |
| --- | --- |
| • Peer-reviewed empirical study.<br>• Published from 2019-2024.<br>• Study involves wearables and/or smartphones for data collection.<br>• Study focuses on modifiable risk factors in metabolic health/disease contexts (minimum one factor from Table S1 is included). | • Article not in English.<br>• Publication type is not an original empirical article (i.e. congress abstract, comments, or letters to the editor).<br>• No quantitative analyses.<br>• Wearable hardware cost ≥500€ (US \$570). |

**Table 2.** Data Charting Form. ^a^

| Charting information | Explanation (if applicable) |
| --- | --- |
| Author(s) |  |
| Year of publication |  |
| Country of origin | Where the study was conducted |
| Population type | Healthy, at-risk, clinically diagnosed |
| Population demographic | Age/Gender/Race |
| Sample size |  |
| Intervention | Yes/No |
| If yes, intervention type |  |
| If yes, duration of intervention |  |
| Type of device | i.e. wrist-worn wearable, mobile application, etc. |
| Modifiable risk factor | i.e. (a) physiological risk factor/ (b) lifestyle risk factor/ (c) or both |
| Modifiable risk factor measure | risk factor type (i.e. physical activity) |
| Unit of measurement | Measure operationalization (i.e., step count per day) |
| Sampling method | i.e. (a) EMA/self-report; (b) passive sensing |
| Measure sampling frequency |  |
| Overall measurement duration |  |
| Number of total risk factors assessed |  |

<sup>a</sup> Note. The data charting form is adapted from prior work [29].

### Stage 5: collating, summarizing, and reporting the results

Following the data charting process outlined in Table 2, the gathered data will be synthesized and summarized in descriptive tables and figures. According to our first research question, we will categorize the modifiable risk factors per each study. Next, we will aggregate the overall prevalence of each modifiable risk factor and the proportion of studies that measure single or multiple modifiable risk factors. Based on our second question, we will describe the measures (or units) used to operationalize the most prevalent factors, and we will overview the range in measurement, i.e., how similarly risk factors are measured across studies. The overall findings will be summarized and communicated through tables and figures.

## Discussion

To our knowledge, this scoping review is among the first to comprehensively summarize the current landscape of key modifiable risk factors in digital metabolic health research, particularly how consistently risk factors are measured across studies. This research will contribute to existing knowledge by systematically scoping the variability in wearable and smartphone-based signals, thus paving the way for improved measurement standards and comparability across studies.

This scoping review will have several limitations, including literature solely in English, which leads to language bias, and limited scope to quantitative empirical studies, which may omit more recent qualitative or industry developments in digital metabolic health. Our results will be specific to wearables that are worn on the body (i.e., we will exclude breath analyzers). Furthermore, the lack of consensus on metabolic health [30] may result in diverse definitions of metabolic risk factors, suggesting the possibility of omission of some factors. Nonetheless, based on prior work, we have identified a comprehensive list of various modifiable risk factors in metabolic health, including lifestyle behaviors and physiological markers [31–38].

The results and findings of this study will be disseminated through publication in relevant journals and presentation at conferences. No ethics approval is required for this study as it does not involve conducting trials or collecting primary data.

## Conclusion

Overall, consolidating the current literature will be a valuable contribution to the field, aiding in describing the prevalence of different risk factors and identifying gaps in measurement. This review also presents new opportunities for establishing standards to enhance the replicability and generalizability of measures in digital metabolic health research.

## Data Availability

N/A

## Acknowledgments

VB, TK, and MJ are affiliated with the Centre for Digital Health Interventions, a joint initiative of the Institute for Implementation Science in Health Care, University of Zurich, the Department of Management, Technology, and Economics at ETH Zurich, and the Institute of Technology Management and School of Medicine at the University of St.Gallen, Centre for Digital Health Interventions is funded in part by Mavie Next, an Austrian health care provider, CSS, a Swiss health insurer, and MTIP, a growth equity firm. TK was also a co-founder of Pathmate Technologies, a university spin-off company that creates and delivers digital clinical pathways. However, Mavie Next, CSS, MTIP or Pathmate Technologies were involved in this protocol. All other authors have no conflicting interests.

## Author Contributions

VB and MJ conceived the idea. VB wrote the first version of the manuscript. TK and MJ provided feedback on the manuscript and revised it. TK and MJ provided methodological guidance.

## Conflicts of Interest

None declared.

## Abbreviations

NCD: noncommunicable disease

## Multimedia Appendix 1

**Table S1.** Search Strategy/Queries

| Terms | Number | Syntax |
| --- | --- | --- |
| Wearables, and smartphones | #1 | "digital biomarker" OR "digital biomarkers" OR "wearable" OR "wearable electronic devices" OR "wearables" OR "smartphone" OR "mobilephone" OR "app" OR "apps" OR "application" OR "applications" OR "implantable" OR "mHealth" OR "ecological momentary assessment" OR "EMA" OR "remote monitoring" OR "fitness tracker" OR "fitness trackers" OR "smartwatch" OR "smartwatches" OR "smart ring" OR "smart rings" OR "heart rate monitor" OR "heart rate monitors" OR "glucose monitor" OR "glucose monitors" |
| **Modifiable risk factors** | #2 |  |
| Physiological markers | #2.1 | "weight loss" OR "weight gain" OR "weightloss" OR "weight change" OR "weight reduction" OR "weight management" OR "body mass index" OR "bmi" OR "adipo*" OR "body composition*" OR "body size*" OR "weight measur*" OR "subcutaneous fat*" OR "abdominal fat*" OR "obesity" OR "overweight" OR "Metabolic Syndrome" OR "obes*" OR "Adiposity" OR "Adipos*" OR "Blood Glucose" OR "Glucose Tolerance Test" OR "Resistance, Insulin" OR "hyperinsulinaemia" OR "glucose metabolism" OR "glucose transport" OR "hyperglycemia" OR "Diabetes Mellitus" OR "Cholesterol" OR "Triglycerides" OR "Dyslipidemia" OR "Epicholesterol" OR "HDL" OR "LDL" OR "Triglyceride*" OR "Hyperlipidem*" OR "Lipidem*" OR "Hypertension" OR "Triglycerides" OR "Hypertens*" OR "Prehypertens*" OR "Systolic" OR "Diastolic" |
| Lifestyle behavior | #2.2 |  |
| Nutrition | #2.2.1 | ("Nutrition Assessment" OR "Nutrition Surveys" OR "Diet" OR "food" OR "foods" OR "Nutrition" OR "Eating" OR "Feeding Behavior" OR "feeding" OR "dietary behavior" OR "dietary" OR "food intake" OR "nutrient" OR "nutrients") |
| Physical activity | #2.2.2 | ("exercise" OR "healthy Lifestyle" OR "physical activit*" OR "physical Fitness" OR "health behavio*" OR "fitness" OR "bicycle" OR "stair climb" OR "strength training" OR "resistance training" OR "active travel" OR "active transport" OR "acceleromet*" OR "walking" OR |
|  |  | "sedentary" OR "physical inactivit*" OR "sitting" OR "screen time" OR "screen use" OR "social media" OR "sport" OR "sports" OR "leisure" OR "leisure active*" OR "run*" OR "training" or "trainings" OR "walk*" |
| Stress | #2.2.3 | "mental health" OR "mental disorder" OR "psychological distress" OR "mental distress" OR "stress" OR "distress" |
| Sleep | #2.2.4 | "sleep*" |
| Substance use | #2.2.5 | "substance" OR "alcohol" OR "drug" OR "opioid" OR "tobacco") AND ("abuse" OR "misuse" OR "withdrawal" OR "abstinence" OR "addict*" OR "craving" OR "dependency" OR "illegal" OR "illicit" OR "overdose" OR "prevention" OR "user" OR "intake" OR "recreational" OR "recovery" OR "smoke" OR "smok*" OR "alcohol*" |
| Publication date | #3 | "2019/01/01"[Date - Publication]: "2024/XX/XX"[Date - Publication] |
| Search query | #1 AND<br>(#2.1 OR<br>#2.2.1<br>OR<br>#2.2.2<br>OR<br>#2.2.3<br>OR<br>#2.2.4<br>OR<br>#2.2.5)<br>AND #3 |  |

## Notes

### Competing Interest Statement

The authors have declared no competing interest.

